# A novel germline EGFR variant p.R831H causes predisposition to familial CDK12-mutant prostate cancer with tandem duplicator phenotype

**DOI:** 10.1101/2020.02.23.20027045

**Authors:** Kaiyu Qian, Gang Wang, Lingao Ju, Jiyan Liu, Hang Zheng, Yingao Zhang, Liang Chen, Yaoyi Xiong, Yongwen Luo, Yejinpeng Wang, Tianchen Peng, Fangjin Chen, Dongmei Liu, Xuefeng Liu, Yi Zhang, Yu Xiao, Xinghuan Wang

## Abstract

5-10% of total prostate cancer (PCa) cases are hereditary. Particularly, immunocheckpoint inhibitor-sensitive tandem duplicator phenotype (TDP) accounts for 6.9% PCa cases, whereas genetic susceptibility genes remain completely unknown. We identified a Chinese family with two PCa patients, in which the PCa phenotype co-segregated with a rare germline variant EGFR^R831H^. Patient-derived conditionally reprogrammed cells (CRC) exhibited increased EGFR and AKT phosphorylation, and a sensitivity to EGFR antagonist Afatinib in migration assays, suggesting the EGFR allele was constitutively active. Both EGFR^R831H^-mutant tumors contained biallelic CDK12 inactivation, together with prominent tandem duplication across the genome. These somatic mutations could be detected in urine before surgery. Analysis of public databases showed a significant correlation between mutation status of EGFR and CDK12. Taken together, our genetic and functional analyses identified a previously undescribed link between EGFR and prostate cancer.

## Introduction

Prostate cancer (PCa) is the most common type of cancer with approximately 449,800 new cases each year. An estimated 6.9% of PCa exhibits biallelic CDK12 inactivation mediated tandem duplicator phenotype (TDP) and is sensitive to immunocheckpoint inhibitor treatment.^1^ A higher frequency of CDK12-truncating mutation associated with TDP has been reported in African Caribbean vs. French Caucasian PCa patients,^2^ suggesting a possible hereditary factor.

Previous studies have reported that approximately 20% of men diagnosed with PCa have a positive family history and that familial risk for first-degree male relatives with PCa is about 2-4 times higher than that in the general population.^3,4^ Though a significant proportion of PCa patients has a positive family history and genome-wide association studies (GWAS) have reported multiple genetic predisposition loci associated with increasd PCa risk, collectively, these fundings only explain a fraction of PCa cases.^5^ BRCA2,^6^ PALB2,^7^ and ATM^8^ are well-documented to cause familial PCa, whereas no application for non-invasive PCa diagnosis has been reported. Besides genes involved in double-stranded break repair, some other hereditary oncogenes, such as MSH2, MSH6, and CHEK2, have been reported in PCa to a lesser extent.^9^ Notably, all reported causal germline variants of PCa reside on DNA-repair-related genes.

Here, we describe a Chinese family with two PCa patients, in which the PCa phenotype co-segregated with a rare, constitutively active germline variant EGFR^R831H^. Interestingly, both PCa tumors contained biallelic CDK12 inactivation and exhibited TDP phenotype.

## Case Report

### Prostate cancer patients in the pedigree

A 64-year-old man (ID: 3557, the proband, Fig S1) presented at our department in Wuhan, China, due to concerns about his family PCa history. Less than a month ago, his brother (ID: 3558) noticed gross hematuria and underwent cystoscopy at our department, revealing a posterior urethral neoplasm extended to bladder (Fig S2). Transurethral resection was performed on patient 3558 and histopathological tissue assessment verified the diagnosis of metastatic PCa. Patient 3558 underwent radical prostatectomy, radical cystectomy, and bladder reconstruction with ileum on November 1, 2018, with a histological confirmation of Gleason score 5+4 (Table S1). Given the patient’s age and family history, a prostate-specific antigen (PSA) test followed by a magnetic resonance imaging (MRI) scan were immediately arranged for patient 3557 (Fig S2-S3). His PSA level was 21.5 ng/mL and MRI scan demonstrated abnormal signals in the peripheral zone of the right lobe using the Prostate Imaging Reporting and Data System (PI-RADS) with score 3. Subsequently, a 13-core prostate biopsy confirmed the PCa diagnosis with Gleason score 4+5 and a radical prostatectomy was performed on December 8, 2018 (Table S1).

### EGFR^R831H^ allele co-segregates with PCa in male members of the pedigree

High throughtput sequencing and health examinations were performed for all members of the family (Table S2-S4). By filtering against the population database, six rare germline mutations were discovered in both 3557 (proband) and 3558. Intersecting the two patients’ germline variants and removing the ones found in known non-PCa male family memberrevealed a single EGFR^R831H^-variant (Fig 1B), with a very low population frequency (0.004%, 13/276986) in the gnomAD global population and 0% (0/18862) in the gnomAD East Asian population. p.R831H is located in the kinase domain of EGFR and found to be a somatic mutation in the public tumor genome databases (COSMIC v88) and a possible germline predisposing mutation in two lung cancer patients.^10^ Primary tumor tissue sequencing revealed an imbalanced copy number gain of the EGFR^R831H^ allele in both tumors, further suggesting that the allele might bring selective advantage to the tumors during their independent evolution. The sole female in the generation and 3557’s older sister, 4351 also had an EGFR^R831H^-mutation. In contrast, the older brother (4350) and the son (4356) of 3557 exhibited no EGFR-mutations with normal PSA levels. In addition, the other four female members (4352, 4353, 4354, and 4355) in the family had no EGFR -mutations and their health examinations reported no tumors (Table S3).

**Fig 1.**
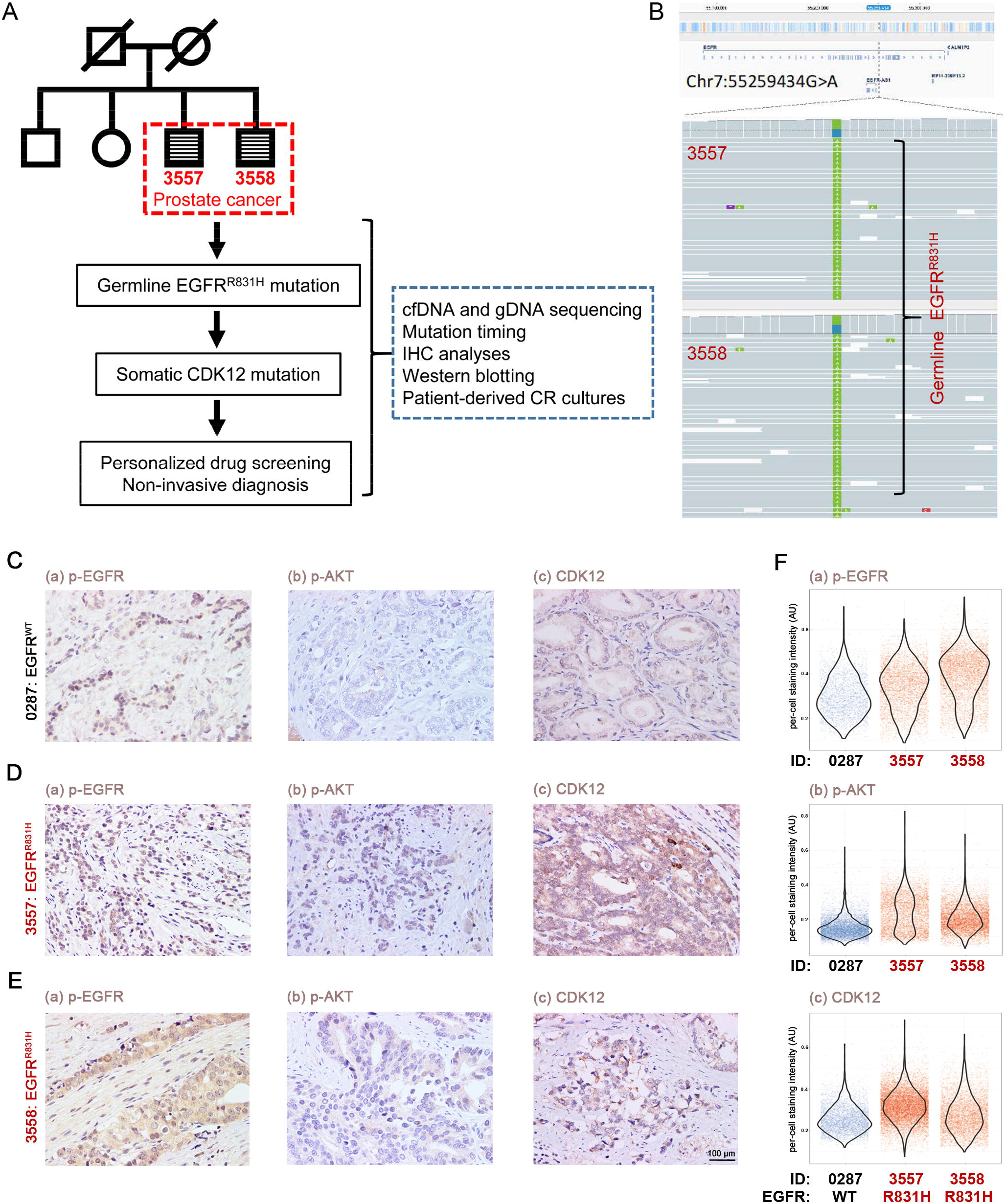
EGFR^R831H^ allele co-segregates with PCa members of the pedigree. **(A)** Overview of the study design. **(B)** Sequencing data revealed mutation 7:55259434 G>A (EGFR^R831H^) in both affected brothers. **(C-E)** IHC stainings for EGFR^WT^ and EGFR^R831H^ PCa tissues showed the expression of p-EGFR, p-AKT and CDK12 in 0287: EGFR^WT^ (C) and EGFR^R831H^ of 3557 (D), 3558 (E). Scale bar: 100 µm. **(F)** Quantification of p-EGFR (a), p-AKT (b) and CDK12 (c) staining intensity indicated increased p-EGFR, p-AKT and CDK12 in 0287 (control, blue) compared to 3557 and 3558 (red). Y-axis represents per-cell staining intensity (AU). Each dot represented a quantified cell and the violin plot shows aggregated distribution of staining intensity.

### Cancer cells expressing EGFR^R831H^ showed enhanced phosphorylation of EGFR and AKT, as well as upregulation of CDK12

Enhanced phosphorylated EGFR (p-EGFR) immunohistochemistry (IHC) staining was present in tumor tissues of 3557 and 3558 compared to the EGFR^WT^ samples (Fig 1C-F, Table S5). Furthermore, the activity of AKT signaling was enhanced in the tumors as evidenced by elevated phosphorylated AKT (p-AKT) staining using IHC (Fig 1C-F, Fig S4) and in EGFR^R831H^ CRCs using Western blot analysis (Fig 2C and 2E, Fig S5), suggesting over-activated downstream signaling of EGFR. Interestingly, expression of CDK12 was also elevated in the EGFR^R831H^ CRCs (Fig 2C and 2E, Fig S5).

**Fig 2.**
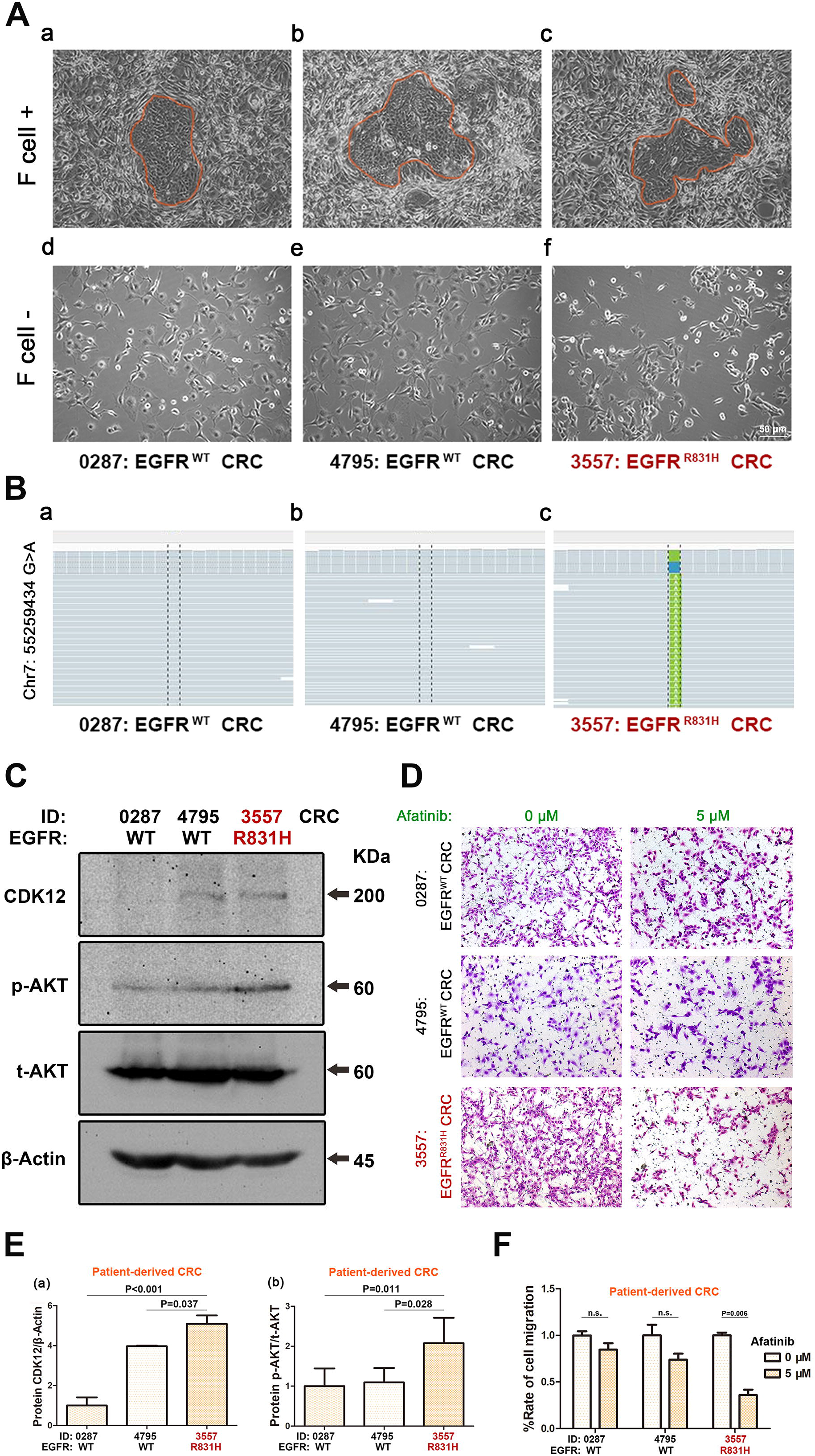
CRCs carrying EGFR p.R831H showed enhanced EGFR signaling as well as upregulation of CDK12. (**A**) Morphology of 0287: EGFR^WT^ CRCs (a and d), 4795: EGFR^WT^ CRCs (b and e) and 3557: EGFR^R831H^ CRCs (c and f) cultured with or without Swiss-3T3-J2 mouse fibroblast feeder cells were observed using phase contrast microscopy. Scale bar is 50 μm. (**B**) Sequencing results showed EGFR^R831H^-mutation only in cell line derived from 3557 tumor.(**C**) Western blot analysis of CDK12 and p-AKT using total protein isolated from patient-derived CRC lysates: 0287 and 4795 (EGFR^WT^ control, black), 3557 (EGFR^R831H^, red). A total of 20□μg protein were loaded per lane. β-Actin and t-AKT were used as loading controls. (**D**) Transwell assay for the CRCs (0287: EGFR^WT^ CRCs, 4795: EGFR^WT^ CRCs, and 3557: EGFR^R831H^ CRCs) pre-treated by Afatinib at 0 and 5 μM for 48 h. **(E)** Protein abundance Western blot analysis of CDK12 (a), p-AKT (b) using total protein isolated from patient-derived CRC lysates: 0287 and 4795 (EGFR^WT^ control, black), 3557 (EGFR^R831H^, red). A total of 20□μg protein were loaded per lane. β-Actin and t-AKT were used as loading controls. Relative intensity for blots of indicated protein measured by Quantity One software. **(F)** For transwell migration analysis, EGFR^WT^ CRCs (0287 and 4795, black) and EGFR^R831H^ CRCs (3557, red) were pretreated by Afatinib at 0 and 5□μM concentrations for 48□h, then incubated in the upper transwell chambers for 24□h. The number of migrated cells was counted in three random fields per chamber using phase contrast microscopy and statistically analyzed.

### CRCs carrying EGFR p.R831H showed enhanced EGFR signaling

Primary cell culture from tumors were derived with the conditioned programmed cell technology (CRC). Western blot results showed that p-EGFR and p-Akt were increased in EGFR^R831H^ CRCs (Fig 2C and 2E, Fig S5), suggesting over-activated downstream signaling of EGFR.

CRC migration rates (0287 and 4795: EGFR^WT^; 3557: EGFR^R831H^) were calculated using transwell chamber migration assays. Compared to other EGFR^WT^ CRCs, the migration rates of 3557: EGFR^R831H^ CRCs were significantly decreased (p=0.006) after a 48□h treatment with Afatinib, further suggesting the that EGFR^R831H^ allele was constitutively active (Fig 2D and 2F). In addition, the migration ability of 3557: EGFR^R831H^ CRCs was stronger than that of 0287 and 4795: EGFR^WT^ CRCs (Fig S6A-B). A similar result was observed in PCa cell line Du145 with EGFR^R831H^ over-expression (Fig S6C-D).

### Biallelic CDK12-mutation and tandem duplicator phenotype in both EGFR-mutant tumors

It was hypothesized that due to similar genetic predisposition the evolution pathway of both EGFR-mutant tumors would be similar. In contrast to the majority of PCa genomes that show chromoplexy-driven punctuative evolution, tumor tissue whole genome and panel sequencing within the family showed no chromoplexy but striking, prevalent, genome-wide tandem duplication of genomic segments (Fig 3A-B), which resulted in oscillating, short-spanned copy number variation (Fig 3C-D).

**Fig 3.**
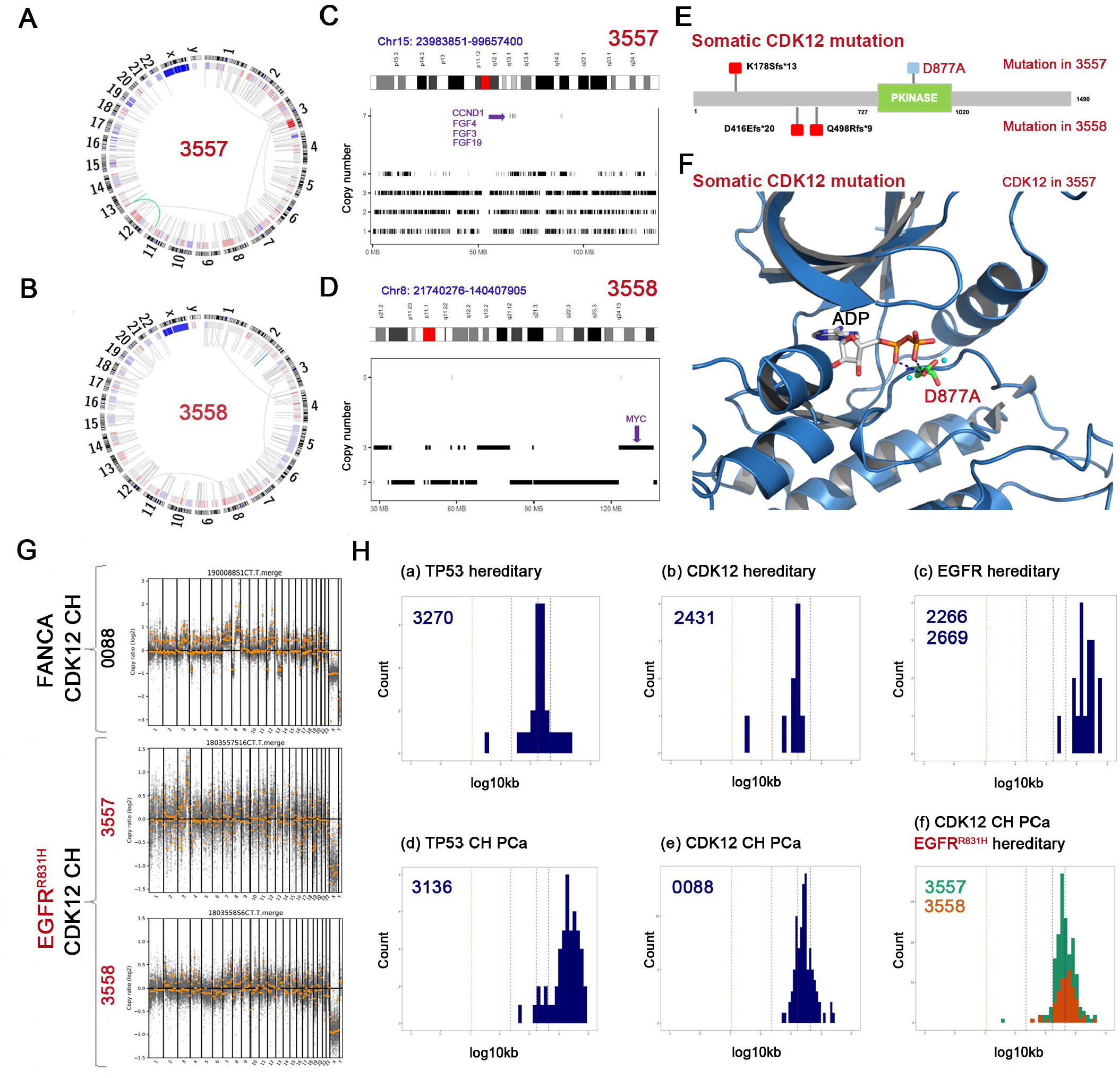
Biallelic CDK12-loss correlates with prominent tandem duplication across the genome. **(A-B)** Circos plot showing prevalent local structural variation (links) across tumor genomes of 3557 (A) and 3558 (B). Structural variations are linked with locally clustered genomic amplifications (red in the external circle) but not deletions (blue in the external circle). **(C-D)** TDP phenotype is prevalent across the genome and amplifies essential oncogenes, such as CCND1/FGF4/FGF3/FGF19 in 3557 chromosome 15 and MYC in 3558 chromosome 8. **(E)** Biallelic CDK12 somatic inactivating mutation found in both tumors. Frameshift mutations were flagged as red mutations, and missense mutation D877A in 3557 was flagged as blue mutation. **(F)** D877A-mutation affects essential residue in ATP-binding-pocket of CDK12 and is predicted as kinase-dead mutation. **(G)** Visually similar pan-genome copy number profile (yellow) with sequencing depth of probe region (gray) generated by CNV kit from 3557 (red), 3558 (red), and 0088 (control, FANCC germline mutation with somatic biallelic CDK12-mutation) showed the characteristic tandem genome duplication phenotype. Hereafter, biallelic mutation is denoted as as compound heterozygous (CH) for clarity. **(H)** Distribution of tandem duplicated genome segment length (on log10 kb scale) in (f) 3557 (green) and 3558 (orange), other tumor samples of (a) TP53 hereditary cancer, (b) hereditary CDK12 rare mutation, (c) hereditary EGFR-mutation, (d) PCa patient 3136 with TP53 biallelic inactivation, and (e) PCa patient 0088 with CDK12 biallelic mutation. Segment length distribution of 3557 and 3558 are highly similar and only similar to 0088 (another CDK12 biallelic mutation tumor) but not to other tumors with EGFR hereditary, CDK12 monoallelic, or TP53 mutations.

TDP is known to be associated with the TP53/BRCA1-mutation, CCNE1 activation, or biallelic CDK12-mutation.^1^ Biallelic CDK12 somatic mutations (three frameshift mutations and one missense mutation, rather than TP53/BRCA1- or CCNE1-mutation) were found in both tumors (Table S2, Fig 3G-H). Besides the three frameshift protein truncating mutations, the only missense mutation (D877A) affecting a critical residue interacting with the ATP/ADP substrate and Mg^2+^ cofactors in the CDK12 kinase domain (Fig 3E-F), suggesting that it is a kinase-dead mutation. Hence, it was concluded that both tumors develop biallelic CDK12 inactivation.

Tandem duplication segment lengths were compared in the EGFR^R831H^/CDK12 biallelic inactivated tumors to other TDP or non-TDP tumors in a pool of in-house tumor samples. As expected, the duplication segment lengths of EGFR^R831H^/CDK12 biallelic inactivated tumors were similar to other CDK12 biallelic inactivated tumors but unlike TP53 biallelic inactivated TDP tumors or other hereditary EGFR-mutant bearing tumors, suggesting that the TDP phenotype was a result of the biallelic CDK12 inactivation (Fig 3H, Fig S7).

### Detection of tumor-specific CDK12 mutations and CNV in urine supernatant

In the proband (3557), both the frameshift (K178SfsTer13) and somatic kinase-dead mutation (D877A) of CDK12 were detected in the cfDNA from urine supernatant before surgery, with 4.23-4.6% variant allele frequency (VAF) (Fig 4B), correlated with the VAF (32-36%) from gDNA of the TDP-PCa tissue sample (Fig 4D). Besides somatic SNV mutation, CNV were also evident in the urine supernatant sample. In contrast, cfDNA from his blood sample before surgery and urine supernatant after surgery showed no somatic CDK12-mutations (Fig 4A and 4C).

**Fig 4.**
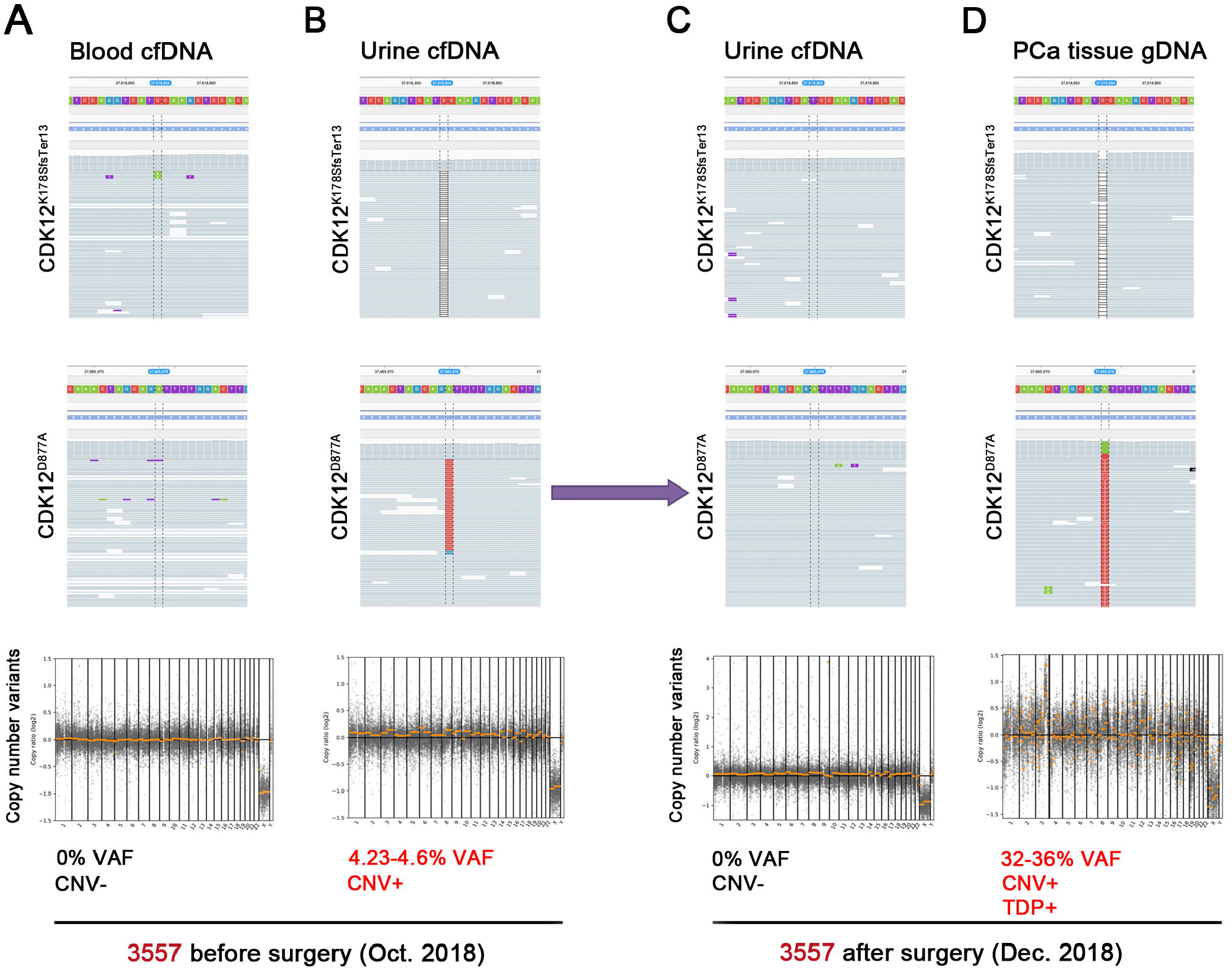
Non-invasive detection of CDK12^D877A^-mutation in urine supernatant. The cfDNA from **(A)** blood and **(B-C)** urine supernanant, **(D)** gDNA from PCa tissue were analyzed in the proband (3557) before and/or after surgery. Both frameshift (K178SfsTer13) and somatic kinase-dead mutation (D877A) of CDK12 were detected in (B) cfDNA from urine before surgery (4.23-4.6% VAF) and (D) gDNA from PCa tissue (32-36% VAF). CNV were also evident in B and D. The tandem duplicator phenotype (TDP) occurs only in the PCa tissue (D).

### Co-occuring EGFR and CDK12 mutants in public tumor databases

To investigate the correlation between EGFR and CDK12 mutations in tumors, EGFR-CDK12 co-occurrences were analyzed in the TCGA and COSMIC databases. The results revealed co-occurrences between EGFR and CDK12 mutations in both databases (Table S6). Furthermore, co-occurring EGFR and CDK12 mutants were analyzed in each tumor type. The EGFR and CDK12 mutations showed significant correlations in certain tumor types from the databases. Additionaly, one tumor sample harboring both EGFR^R831H^- and CDK12-mutations also processed a CDK12-mutation (Table S7). These results suggest that EGFR mutation might predispose a specific vulnerability to CDK12 somatic mutation. Hereditary mutations of EGFR have been previously reported to account for lung cancer (p.T790M,^11^ p.V843I,^12^ and p.V834L^13^). However, a link with PCa has yet to be determined. None of the reports^10^ described rare EGFR variant carriers with prostate cancer. However, one study^13^ described a pedigree with EGFR p.V834L co-segregates with lung cancer, which contains a prostate cancer patient with unknown germline mutation status.

## Discussion

In this study, a rare germline EGFR^R831H^-variant was identified in members of a Chinese family affected by TDP-PCa. Pathogenicity of the EGFR^R831H^ allele was supported by multiple lines of experimental and statistical investigations: rarity in population, co-segregation of PCa phenotype in male family members, strategic positioning in the kinase domain of EGFR, selective, allelei-specific copy number gain in tumor tissues, enhanced *in vitro* and *in vivo* EGFR downstream signaling activity, and decreased EGFR^R831H^-mutant CRC migration under selective EGFR pharmacological inhibition. In addition, development of a rare molecular subtype PCa in different affected members with functionally similar somatic mutation profiles further suggested the tumors had a genetic predisposition. Hence, it was concluded that the germline EGFR^R831H^-variant predisposes to TDP-PCa in this family.

Biallelic CDK12 somatic mutations were found in both tumors, including three frame-shift, likely protein-null mutants, and one likely kinase-dead mutant (D877A) in the ATP-binding pocket. The somatic CDK12 mutations were detected in the cfDNA from the urine of patient 3557 before surgery, indicating a potential application for non-invasive diagnosis and monitoring for the TDP-PCa. All CDK12-mutations were clonal in the tumors, suggesting that they were among the earliest tumor evolution events. Besides the CDK12-mutations, a few passenger somatic short-nucleic-acid-variation (SNV) mutations and no potential driver mutations were identified in the tumors. These results collectively suggest that biallelic CDK12 inactivation is necessary and sufficient to drive oncogenesis in these tumors.

Biallelic CDK12 inactivation was documented for 6.9% of metastatic castration-resistant PCa types in a previous study.^1^ It was mutually exclusive to other known molecular subtypes of PCa, including ETS fusion, dMMR, and SPOP-mutation. PCa cases with biallelic CDK12 inactivation were mostly metastatic, and characterized by specific tandem genomic duplication phenotypes^1,14^ and specific susceptibility to immunocheckpoint therapy.^1^ Considering the chance probability of both patients in this family developing a biallelic CDK12 inactivation subtype of PCa is 0.461%, it is unlikely to happen by mere chance and is likely to be influenced by specific environmental factors or hereditary predisposition. Taken together, these results supported the EGFR^R831H^ allele as a hereditary germline predisposing variant.

A following question is how their common genetic predisposition drives similar somatic evolution, namely, how constitutive EGFR activation leads to adaptive CDK12 inactivation. CDK12 protein level was found to be increased in tumor IHC staining *in vivo* and in CRCs *ex vivo*, suggesting that CDK12 expression level might be elevated under constitutive EGFR activation. Data mining from public tumor sequencing databases showed a correlation between EGFR and CDK12 mutations. Furthermore, an additional tumor carrying the EGFR^R831H^ allele and CDK12-mutation were documented in the COSMIC database. Additional studies are needed to characterize this molecular mechanism in detail.

Constitutively active EGFR mutations have been well documented in other cancer types, particularly lung adenocarcinoma. Few studies, if any, have documented the pathogenic EGFR mutations in PCa. To the best of our knowledge, this study is the first report identifying a pathogenic EGFR germline mutation in PCa.

In conclusion, this study identified a rare germline EGFR^R831H^-variant that predisposes to PCa with a specific molecular subtype characterized by biallelic CDK12 inactivation and tandem genome duplication. Constitutively active EGFR mutations have been well documented in other cancer types, particularly lung adenocarcinoma. Few studies, if any, have documented the pathogenic EGFR mutations in PCa. To the best of our knowledge, this study is the first report identifying a pathogenic EGFR germline mutation in PCa. Our results not only expand the gene list for hereditary PCa but also contribute to documenting the complexity of EGFR mutation pathogenicity. These findings can also suggest venues for germline screening, non-invasive diagnosis, and monitoring in clinical practice, as these tumors might be sensitive to targeted therapy and immunocheckpoint inhibition.

## Methods

### Ethical statement for patients and family members in this study

Clinical, pathological, and follow-up data records for all PCa patients and their family members were collected (Table S1-S2) and two pathologists were invited to independently confirm the histology diagnosis. Additional cancer patient samples were collected through the clinical practice in the Zhongnan Hospital of Wuhan University and West China Hospital of Sichuan University. The study use of clinical information and human samples (including blood, urine supernatant, surgical tissue specimens, and primary cancer cells) was approved by the Ethics Review Committee at Zhongnan Hospital of Wuhan University (approval number: 2015029). Human sample preservation by the Zhongnan Hospital Biobank, the official member of the International Society for Biological and Environmental Repositories (https://irlocator.isber.org/details/60), was approved by the Ethics Review Committee at Zhongnan Hospital of Wuhan University (approval number: 2017038) and China Human Genetic Resources Management Office, Ministry of Science and Technology of China (approval number: 20171793). All PCa patients, family members of the PCa pedigree, and additional cancer patients, provided written informed consent. All study procedures were performed in accordance with the ethical standards of the Institutional Ethics Review Committee.

### DNA sequencing data pre-processing

Raw sequencing data were trimmed for adaptors and homopolymer tails, and controlled for sequencing quality. The cleaned data were mapped to the human genome (GRCh37) with bwa (0.7.9). Duplicate read marking was performed using sambamba (0.5.4). Base-quality recalibration was accomplished with Sentieon (Sentieon-Genomics-201808.05).

Hereditary/somatic tumor mutation analysis is described in Supplement.

### Tandem duplicator phenotype (TDP) determination

Copy number variations in tumors were called by CNV kit (0.9.3). Structural variations were called by iCallSV. Possible chromoplexy events were searched by ChainFinder (1.0.1). Copy number oscillations in concordance with structurally split/discordant reads were computed by ShatterSeek (0.4). An in-house R package was used for visualizing the copy number gain structural variant lengths. In-house samples of known tandem duplicator phenotype and normal tumors were used for comparison.

### Hereditary tumor mutation analysis

Germline tumor mutations were called with the Sentieon haplotyper (Sentieon-Genomics-201808.05) and annotated with VEP (90.1) and SnpSift (4.2). Candidate germline variants were filtered with the gnomAD global frequency <0.001 and in-house database frequency <0.001 (out of 20,000 patients). Intersection with variants found in different male members of the pedigree was performed to extract patient-specific germline mutations.

Detailed information is shown in the Supplementary information.

## Data Availability

The datasets used and/or analysed during the current study are available from the corresponding author on reasonable request.

## List of abbreviations

BAF: B-allele frequency
COSMIC: Catalogue of somatic mutations in cancer
cfDNA: Cell-free DNA
CRC: Conditionally reprogrammed cell
CNV: Copy-number-varied
FFPE: Formalin-fixed paraffin-embedded
GWAS: Genome-wide association studies
IHC: Immunohistochemistry
ORF: Open reading frame
p-AKT: Phosphorylated AKT
p-EGFR: Phosphorylated EGFR
PCa: Prostate cancer
PI-RADS: Prostate imaging reporting and data system
PSA: Prostate-specific antigen
RT: Room temperature
SNV: Short-nucleic-acid-variation
TDP: Tandem duplicator phenotype
TCGA: The cancer genome atlas
VAF: Variant allele frequency

## Authors’ contributions

KQ, GW, LJ, JL, HZ, YZ, ZY, FC, DL, RL, WJ, YG, HM, SL and YZ performed experiments. All authors contributed to the research. KQ, GW, LJ, JL, WJ, YZ, YX and XW designed the study and wrote the manuscript. All authors approved the final manuscript.

## Acknowledgments

We thank the patients and their family members for participating in our study. We grateflully acknowledge excellent technical assistance provided by Ms. Yuan Zhu, Ms. Shanshan Zhang, and Ms. Yayun Fang from Zhongnan Hospital of Wuhan University. We would like to acknowledge the TCGA and COSMIC databases for providing use of data free of charge. We also thank International Science Editing (http://www.internationalscienceediting.com) for editing this manuscript.

